# Decoding Prodromal Parkinson’s Disease: Clinical Differences between Isolated Hyposmia and REM Sleep Behavior Disorder

**DOI:** 10.1101/2024.03.18.24304459

**Authors:** Luke Vikram Banerjee, Jacopo Pasquini, Robin Henderson, Nicola Pavese, Kirstie N Anderson

## Abstract

**Background:** The prodromal phase of Parkinson’s disease (PD), much like the disease itself, displays marked heterogeneity, with varied rates of progression and symptom severities. A detailed clinical characterization of prodromal subgroups may provide useful insights for both clinical and research settings.

**Objectives:** To compare clinical assessments in patients with idiopathic rapid eye movement sleep behavior disorder (iRBD) and those with isolated hyposmia.

**Methods:** A cross-sectional study was conducted on 191 patients with iRBD, 213 patients with isolated hyposmia and 150 healthy controls recruited in the Parkinson’s Progression Markers Initiative. The earliest available assessment for each participant was selected. Our analysis investigated and compared the Montreal Cognitive Assessment, Scales for Outcomes in Parkinson’s Disease Autonomic Dysfunction (SCOPA-AUT) and Movement Disorder Society Unified Parkinson’s Disease Rating Scale (MDS-UPDRS) Parts I, II and III scores across the three groups. To assess differences, after adjusting for age and sex, we employed permutations testing. We further investigated the specific question items that contributed most significantly to the observed variations between the groups.

**Results:** We found significant differences between the healthy control group and a combined prodromal group across all assessment categories, with prodromal participants displaying poorer scores. For between prodromal groups comparison, significant differences emerged in SCOPA-AUT and MDS-UPDRS Part I scores, with the iRBD group presenting with more severe scores.

**Conclusion:** Our study highlights that even in the premotor stage of PD, clinical distinctions exist in terms of autonomic burden between individuals with iRBD and those with isolated hyposmia.

## Introduction

Parkinson’s disease (PD) has emerged as the most rapidly growing neurological disorder. The World Health Organization reports global prevalence has approximately doubled from 1994 to 8.5 million in 2019, accounting for 5.8 million disability-adjusted life years and 329,000 deaths (1). Notably, the rising incidence of PD cannot be solely attributed to the aging global population, as the incidence continues to increase even when adjusted for age (2).

PD features a well-established prodromal phase, where neurodegeneration occurs long before the manifestation of classical motor symptoms (3,4). Both PD and its prodromal phase are highly heterogeneous, marked by differences in the speed of disease progression, age of onset, and non-motor symptoms (5,6). However, there is limited research directly examining the heterogeneity within the prodromal phase itself.

The “Brain-first vs Body-first” hypothesis proposed by Horsager et al., offers one theoretical framework to explain the variability of PD (7). This hypothesis delineates two pathological trajectories for PD: the body-first pathway, where neurodegeneration initiates in the gut and then ascends to the brain, with the nigrostriatal neurons affected at a relatively later stage; and the brain-first pathway, where neurodegeneration begins in the limbic system or olfactory bulb and spreads downwards, with the nigrostriatal neurons affected relatively earlier (5). Idiopathic rapid eye movement sleep behavior disorder (iRBD) preceding motor symptoms is considered a strong indicator of the “body-first” subtype. This subtype is characterized by a more aggressive disease course, marked by rapid progression and severe cognitive and behavioral symptoms. Conversely, the “brain-first” subtype generally lacks iRBD at onset, demonstrates a slower progression, and presents with fewer cognitive and behavioral symptoms. Various data-driven studies have also indicated the presence of multiple subtypes within PD, with the recurrent identification of a ‘diffuse malignant’ subtype featuring iRBD (8–12).

All subtypes are thought to converge to a similar stage with widespread pathological involvement. This makes the period before the onset of motor symptoms a crucial window for distinguishing between prodromal subtypes (13).

Our research focuses on the identification of clinical markers in subjects with iRBD compared to those with isolated olfactory loss, both considered strong risks factors for PD. We aim to provide a broad description of these premotor syndromes by examining clinical assessments of cognitive ability, autonomic dysfunction, behavioral and motor symptoms. This study will consider whether this pattern of clinical scores suggests distinct prodromal phenotypes. Overall, we hope to enrich current understanding of the clinical phenotypes of premotor parkinsonism, which may help guide both clinical and research practices.

## Methods

### Data

Data was acquired from the Parkinson’s Progression Markers Initiative (PPMI) database, downloaded on December 21st, 2023. PPMI is a worldwide, multicenter, longitudinal cohort study which aims to identify biological markers of Parkinson’s disease onset and progression. Data is available at https://www.ppmi-info.org/access-data-specimens/download-data (Research Resource Identifier:SCR_006431). For current study details visit www.ppmi-info.org. This study used tier 1 data, openly available from PPMI, and tier 3 data, obtained from PPMI upon request after approval by the PPMI Data Access Committee.

For each participant assessment the earliest datapoint of either the screening or baseline visit was selected. For participants with missing datapoints, we employed a listwise deletion approach.

### Population

Prodromal participants with iRBD or isolated hyposmia, alongside a group of healthy controls were extracted from the PPMI database.

Our study excluded, for all groups, those with a diagnosis of PD, under the age of 50 years, and carriers of known PD genetic variants (Leucine-Rich Repeat Kinase 2, β-Glucocerebrosidase, Alpha-Synuclein, Parkin RBR E3 Ubiquitin Protein Ligase, and PTEN Induced Putative Kinase 1) (14).

Participants with iRBD were recruited to PPMI with diagnosis confirmed either by polysomnography or clinical history evaluation by a sleep specialist (15). This included the management of any concurrent moderate or severe obstructive sleep apnea, as necessary. Clinically diagnosed iRBD participants without polysomnography confirmation required two additional risk factors to be included: hyposmia and DAT-SPECT that was either visually abnormal or reduced from age expected values as determined by quantitative analysis (16).

Participants with isolated hyposmia were recruited to PPMI if they had a Parkinson’s Associated Risk Syndrome (PARS) score of 9 or below and DAT-SPECT imaging that was either visually abnormal or reduced from age expected values as determined by quantitative analysis (17). The PARS score is derived from an algorithm that includes age, sex, and University of Pennsylvania Smell Identification Test (18). If participants in the database were initially classified only as isolated hyposmia but the prodromal history data revealed diagnosis of iRBD confirmed with polysomnography, we reclassified them into the iRBD group.

DAT-SPECT visual assessment was performed by a nuclear medicine expert at the PPMI imaging core lab located within the Institute of Neurodegenerative Disorders. As per the PPMI protocol, it was estimated that the study would recruit approximately 75% of participants with a DAT deficit, defined by hybrid of visual assessment and quantitative striatal specific binding analysis. The remaining 25% had either a reduction from age-expected values, but not outside the normal range, or possessed specific PD risk factors, such as polysomnography confirmed iRBD or rare genetic variants (as previously stated, rare genetic variants were excluded from this study).

Healthy controls were included based on the following criteria: they had no first-degree relative with a PD diagnosis, no significant neurological disorders, and exhibited a visually normal dopamine transporter scan using single photon emission computed tomography (DAT-SPECT).

Detailed information regarding the inclusion and exclusion criteria for the PPMI study is available on the PPMI website (19).

### Clinical Assessments

Motor and non-motor manifestations’ scores were collected. The Montreal Cognitive Assessment (MoCA) assessment was used to assess cognitive dysfunction (20–23). The Scales for Outcomes in Parkinson’s Disease Autonomic Dysfunction (SCOPA-AUT) was used to quantify the burden of autonomic symptoms (24–26). However, SCOPA-AUT sex-related questions were omitted from our statistical analysis, as a significant proportion of participants across all subgroups selected ‘not applicable’ in response to these items. The Movement Disorder Society Unified Parkinson’s Disease Rating Scale (MDS-UPDRS) parts I, II and III were used to score both non-motor and motor manifestations (27).

### Statistical Analysis

Data was pre-processed using Python 3 (28) and then imported into R Studio for statistical analysis (29). The primary analysis was to assess the mean differences in total scores between groups. Before hypothesis testing, assessments scores were adjusted for the effects of age and sex, as these differed significantly between groups. We adjusted for these confounding variables using a two-stage path analysis (30). In the first stage we constructed a linear model to regress assessment scores on age and sex only, and at the second stage we considered group effects on the model residuals, which were uncorrelated with age and sex. Thus, having explained as much variation as possible using age and sex, we can assess whether any of the residual variation can be explained by differences between groups.

Permutations testing was employed (with 10,000 permutations) to evaluate the statistical significance of the differences observed between groups after adjustment for age and sex (31). This non-parametric approach was chosen because responses were both discrete and skewed, meaning the assumptions of the linear model would be invalid. To compare scores across three groups and to maintain independence, we decided to perform two orthogonal tests. Our first was a comparison between a combined prodromal group (both iRBD and hyposmic participants) against the healthy control group. Our second was a comparison between the two prodromal groups. Statistical significance was determined using a predefined threshold: a p-value below 0.05.

Finally, means and 95% confidence intervals for the differences in individual question responses within assessments were plotted. This approach was designed to highlight specific questions that contributed most significantly to the variance observed in the overall assessment outcomes. Hypothesis testing was not conducted for individual scales’ subitems because it was not the main purpose of the analysis, and a multitude of statistical tests would have resulted from this approach.

## Results

List-wise deletion resulted in the exclusion of 27 participants with incomplete data: seven from the iRBD group, eighteen from the hyposmic group, and two from the healthy control group. One participant from the iRBD group was excluded due to the use of dopaminergic medications, rendering the MDS-UPDRS Part III non-comparable. Following these exclusions, a total of 554 participants were included for full analysis (Table 1). The iRBD group was significantly older, followed by the hyposmic group, with the healthy group being the youngest. The iRBD group was predominately male, in contrast to the hyposmic group which showed a higher proportion of females, as detailed in Table 1. This gender distribution aligns with well-documented evidence indicating a higher prevalence of iRBD among males compared to females (32). Within the iRBD group, 133 participants had polysomnography confirmed iRBD, while the remaining 58 were identified based on clinical history (with abnormal DAT-SPECT and hyposmia). Table 1 and Figure 1 present the DAT-SPECT striatal binding ratio values, highlighting anticipated disparities between the healthy control group and the two prodromal groups. In these comparisons, the healthy controls exhibit higher scores relative to both prodromal groups.

**Figure 1:**
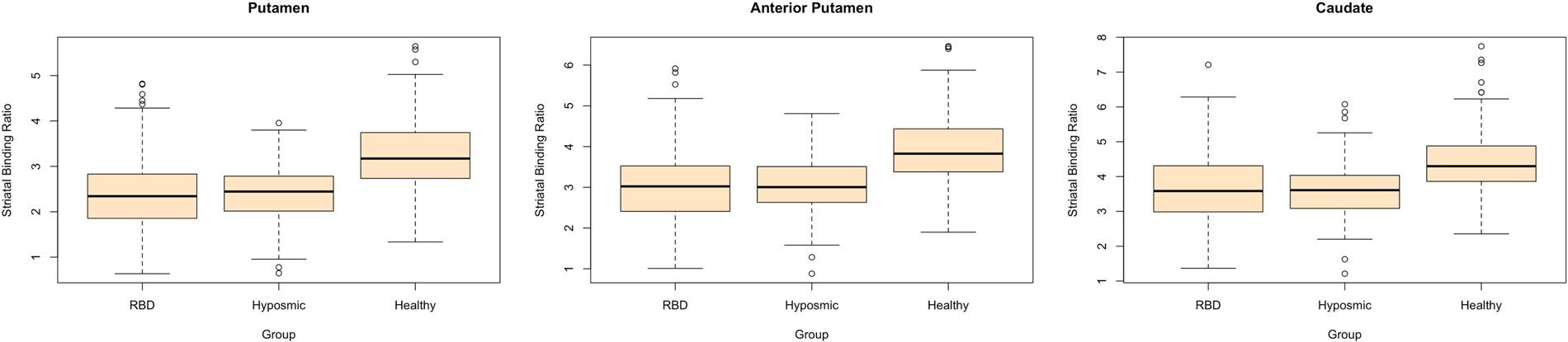
Boxplots illustrate variations in DAT-SPECT striatal binding ratios among different groups, segmented by brain region, with the occipital cortex serving as reference tissue (16). Data were missing for one iRBD participant and four healthy controls.

**Table 1.**
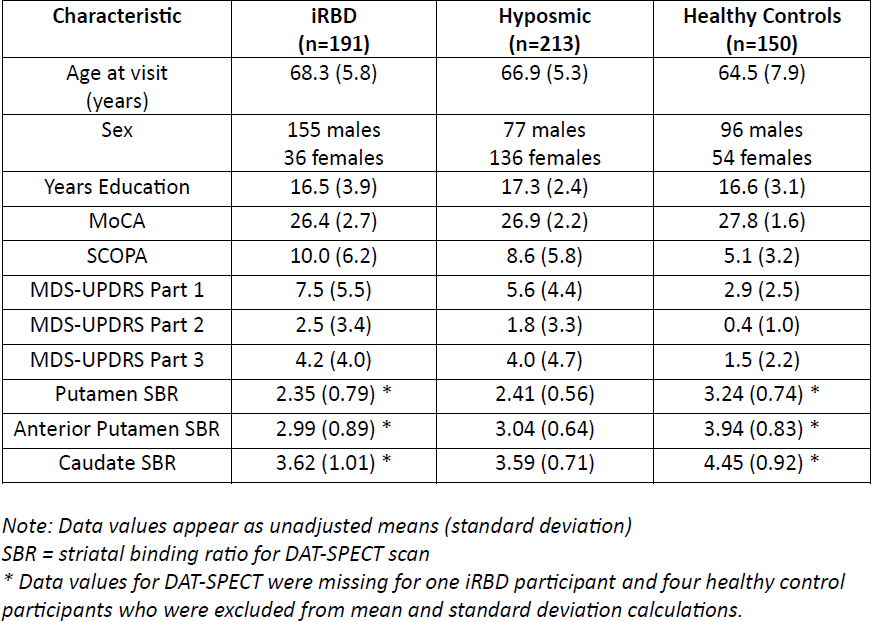
Participant Characteristics.

The prodromal combined group exhibited statistically significant differences across all variables when compared to the healthy control group (all p-values < 0.0001, Figure 2). This was characterized by a lower mean MoCA score and higher mean scores MDS-UPDRS parts I, II and III, and for SCOPA-AUT. The comparison between the iRBD group and the hyposmic group revealed statistically significant higher scores for the iRBD group in the SCOPA-AUT and MDS-UPDRS Part I, with p-values 0.0022 and < 0.0001, respectively. No significant differences between the iRBD group and hyposmic group were observed in the MoCA, MDS-UPDRS Part II, and MDS-UPDRS Part III scores.

**Figure 2:**
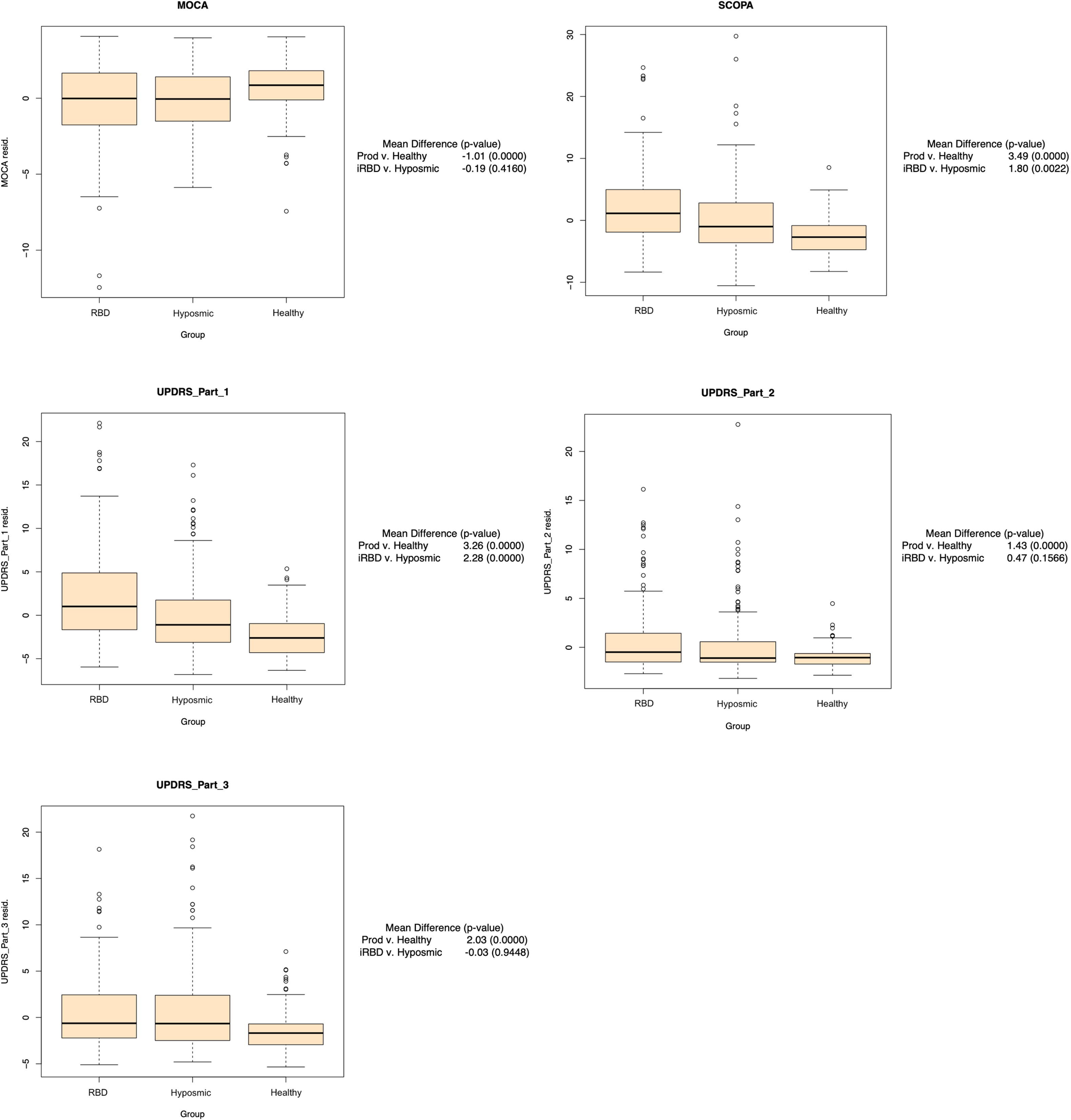
Boxplots display residuals for variables by group. Residuals were derived from linear regression with age and sex as covariates. This highlights differences between groups after adjusting for age and sex.

To investigate the specific questions that contributed the most to overall between-group differences, the mean differences for individual questions with 95% confidence intervals were plotted in Figure 3. Adjustments for age and sex do not alter the ranking of these contributions. In the comparison between healthy controls and the prodromal combined group, the most pronounced individual question differences were as follows: for the MoCA memory recall questions were the most significant; for SCOPA-AUT, straining to pass stools, constipation, and nocturia; for MDS-UPDRS Part I, sleep problems at night, pain, fatigue and constipation; for Part II, handwriting, tremor, and salivation; and for Part III, left finger tapping, left toe tapping, and kinetic tremor in the left hand, with the prodromal group consistently showing greater symptom severity compared to the healthy group. Where significant differences were observed between prodromal groups, the most significant individual questions were for SCOPA-AUT, straining to pass stools, constipation, and a weak urine stream; and for MDS-UPDRS Part I, daytime sleepiness, depression, and constipation, with the iRBD group consistently showing greater symptom severity than the hyposmic group.

**Figure 3:**
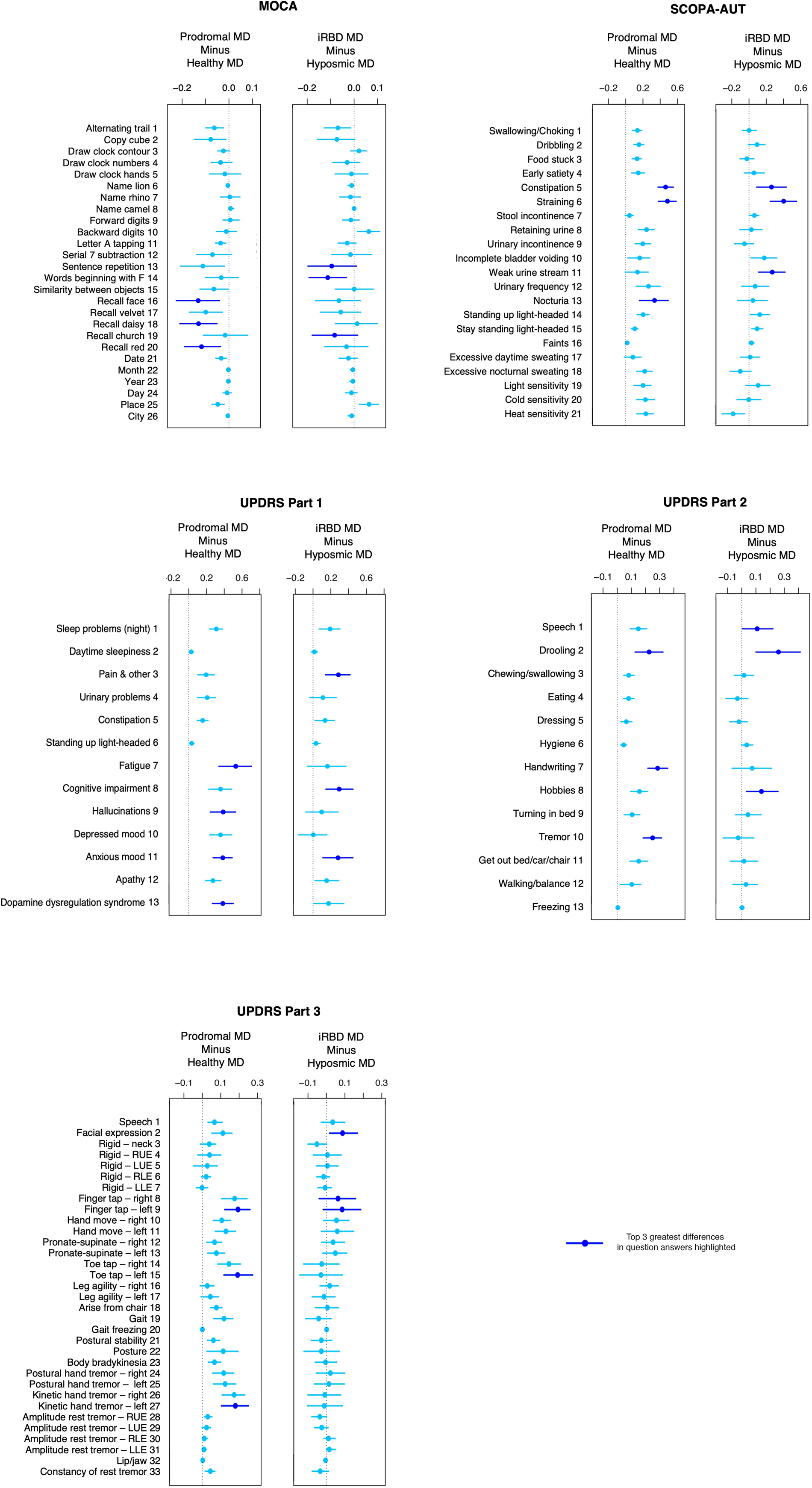
Differences in question answers between groups. Questions are on the y-axis and mean difference (MD) with 95% confidence intervals on the x-axis. For each assessment, left shows: prodromal mean minus healthy mean, right shows: iRBD mean minus hyposmic mean.

## Discussion

Our results reveal early clinical differences between the two proposed premotor phenotypes of PD, marking one of the largest prodromal cohort studies to date. These findings provide new evidence supporting the existence of distinct subgroups of PD in the prodromal phase. We also identify early clinical differences between prodromal participants and healthy controls.

There are limited studies characterizing the subtypes of PD in the prodromal phase, in part due to the challenge of appropriately identifying and recruiting participants. While iRBD is a well-recognized and robust clinical biomarker for prodromal PD, isolated hyposmia shows considerably lower specificity for PD (3,33,34). Previous studies on the prodromal phase have focused only on individuals with iRBD, omitting those without iRBD (35,36). This approach misses a significant subset of PD patients who will never have iRBD, thereby limiting a comprehensive understanding of the prodromal phase (37,38). Our study addresses this gap by incorporating a hyposmic cohort, enriched by the prerequisite of abnormal DAT-SPECT imaging for enrolment.

The comparison between groups shows an overall deterioration in all motor and non-motor assessments considered in this study in the combined prodromal group relative to healthy controls. This observation provides additional evidence for a progressive involvement of the central nervous system in PD.

In the comparison between the iRBD and hyposmic groups, there were significant differences in the SCOPA-AUT and MDS-UPDRS Part I scores. Given the substantial overlap in question content of SCOPA-AUT and MDS-UPDRS Part I, the concurrence of findings is expected. These findings demonstrate that participants in the iRBD group have significantly greater burden of autonomic symptoms when compared to the hyposmic group, lending support to the idea that iRBD patients and PD patients with RBD have greater peripheral nervous system involvement at an earlier stage. This broadly aligns with the Brain-first vs Body-first hypothesis (5). However, the hyposmic group also displayed elevated autonomic scores compared to healthy controls, albeit less pronounced than in iRBD cases. This could suggest hyposmic individuals have peripheral nervous system involvement prior to motor symptoms. Though, this could also be explained by central mechanisms, with hypothalamic involvement causing autonomic dysfunction. When interpreting these findings, it must be noted that the PPMI study targeted a prodromal cohort enriched with a DAT deficit. Despite this, the emergence of autonomic symptoms in hyposmic participants, either before or with motor symptoms, is significant.

Analysis of the individual question differences revealed constipation as a consistent theme accounting for the greater scores in the iRBD group compared to hyposmic. This could indicate a higher diagnostic value for constipation in PD in the context of iRBD patients compared to those only with hyposmia. The absence of differences between iRBD and hypsomic groups in MoCA, MDS-UPDRS Part II and Part III scores at this stage could be attributed to the disease not having progressed enough to manifest significant cognitive or motor symptoms in either group.

A notable limitation of our study’s methodology is the assumption that the iRBD and hyposmic subgroups are at comparable stages of disease progression. The average duration until phenoconversion could vary significantly between prodromal groups, with some participants potentially never undergoing phenoconversion. Future studies could mitigate this limitation by using the PPMI dataset, which gathers longitudinal data on participants. This would allow for retrospective analyses at equivalent time points before phenoconversion, enabling a more accurate comparison between groups. However, the broad equivalence of the DAT-SPECT striatal binding ratio of the prodromal groups, coupled with their proven predictive value for phenoconversion, suggests that the effect of this limitation might be minimal (39,40).

Our study operates under the assumption that there are only two prodromal phenotypes of PD, potentially overlooking additional phenotypic variations. Despite this limitation, our research could still contribute valuable insights by establishing broad phenotypic classifications, within which specific subgroups can be identified and further studied.

An additional limitation of our study is the cross-sectional nature of our analysis, focusing on assessments at a single time point. Consequently, we have not accounted for the evolution of clinical assessments over time between groups. The completion of the PPMI dataset will enable such longitudinal studies, offering insights into how these clinical markers progress and diverge between groups over time.

Our study’s participant pool was drawn exclusively from the PPMI dataset and excluded known genetic PD variant carriers. This may not capture the full spectrum of prodromal PD, especially those that do not meet the dataset’s inclusion criteria, such as those with rapidly progressing phenotypes that may be overlooked due to their swift symptom evolution and early phenoconversion.

In conclusion, our study has provided additional evidence supporting the existence of two clinically distinct phenotypes within the prodromal phase of PD. Future research is essential to validate these preliminary findings through longitudinal studies and by exploring additional dimensions, including imaging and various biomarkers.

## Acknowledgements

We would like to thank the study participants who generously contributed their time and effort. The Michael J. Fox Foundation and Parkinson’s Progression Markers Initiative for their support and generation of the database. It is anticipated that the Aligning Science Across Parkinson’s organisation may provide open access funding, contingent upon its acceptance. Confirmation of the funding support will be updated in the final version of this manuscript. We also wish to thank the staff at the Clinical Ageing Research Unit, Newcastle University, for their commitment to this research. The ChatGPT 4.0 large language model provided writing assistance by suggesting alternative phrasings for small sections of text originally written by the authors, it was used for no further purposes.

## Author Roles

Luke Vikram Banerjee – design, execution, data analysis, writing, editing of final version of the manuscript.

Jacopo Pasquini – design, execution, writing, editing of final version of the manuscript, subject matter expert.

Robin Henderson – statistician, data analysis, writing.

Nicola Pavese – execution, writing, editing of final version of the manuscript, subject matter expert.

Kirstie N Anderson – execution, writing, editing of final version of the manuscript, subject matter expert.

## Financial Disclosures

### Relating to current research covered in this study regardless of date

LVB, JP and RH report no financial disclosures or conflicts of interest in relation to the manuscript. NP and KNA receive grant funding from the Michael J Fox Foundation to run the Parkinson’s Progression Markers Initiative study in Newcastle upon Tyne.

### All disclosures for preceding 12-months regardless of relation to this study

LVB, JP and RH report no sources of financial support and funding for the past year, including those unrelated to the manuscript. Author NP sits on advisory boards for Bial, Hoffmann-La Roche, Inc., and Abbvie, and receives grants from the Independent Research Fund Denmark, Danish Parkinson’s disease Association, Parkinson’s UK, GE Healthcare Grant, Multiple System Atrophy Trust and F. Hoffmann-La Roche, Inc. Author KA has attended advisory boards for Janssen Cilag, Idorsia and has received grant funding from Closing the Gap mental health network, and has had speakers fees from Bioprojet, Flynn Pharma and Idorsia.

